# The Economic Burden of Bronchiolitis Obliterans Syndrome After Allogeneic Hematopoietic Cell Transplantation

**DOI:** 10.64898/2026.11.20.26368587

**Authors:** Abdelrahman M. Attia, Adarsh Suresh, Raj Patel, Hugo Escobido Lara, Curtis Lee, Hannah Smati, Ryan Ramphul, Suja Rajan, Lara Bashoura, Burton F. Dickey, Bina Patel, Gabriela Rondon, George L. Chen, Portia Smallbone, Uday R. Popat, Chitra Hosing, Partow Kebriaei, Richard E. Champlin, Elizabeth J. Shpall, Amin M. Alousi, Hemang Yadav, Guang-Shing Cheng, Daniel Wolff, Ajay Sheshadri

## Abstract

**Research Question:** Among adult first allogeneic HCT recipients sampled in a nested case-control study, how did bronchiolitis obliterans syndrome (BOS) affect institutional cost trajectories compared to extrapulmonary chronic graft-versus-host disease (cGVHD) and no cGVHD, and can this information guide cost-effective implementation of home spirometry?

**Study Design and Methods:** We linked clinical, pulmonary function test (PFT), vital-status, and institutional billing data for 308 patients from a nested case-control study: 111 with BOS, 132 with cGVHD without BOS, and 65 without cGVHD. Comparator patients were matched to BOS cases by age (within 10 years), transplantation era (5-year intervals), and time since HCT (within 1 month). Monthly costs from M-6 through M+6 were standardized to 2024 US dollars using CPI-U Medical Care. Gaussian identity-link generalized estimating equations (GEE) modeled clinical group, month, and the group-by-month interaction to calculate within-group pre/post and difference-in-differences (DiD) tests. BOS surveillance analyses evaluated clinic PFT frequency, FEV₁ trajectories, and home-spirometry break-even and net monetary benefit (NMB) scenarios, incorporating projected monitoring costs, avoided BOS-associated costs, and the monetary value of health benefits.

**Results:** Mean monthly cost fell after index in patients without cGVHD by $7,582 (95% CI, -$13,378 to -$1,786; p = 0.01), but not in cGVHD (-$5,768; p = 0.08) or BOS (+$4,470; p = 0.23). The BOS trajectory was $12,052/month less favorable than no cGVHD (95% CI, $2,700-$21,404; p = 0.01), a six-month differential of $72,313. The BOS-vs-cGVHD DiD was $10,238/month in BOS (p = 0.04). Antecedent decline was documented in 40/108 BOS cases (37.0%), a median of 268 days before diagnosis. Each additional clinic PFT/year was associated with higher detection odds (OR, 2.38; 95% CI, 1.41-4.02; p = 0.001). Among BOS cases in the parent cohort, undetected impairment was not significantly associated with geographic proximity to comprehensive cancer centers or socioeconomic vulnerability. At an assumed BOS frequency of 5%, targeted home monitoring would need to prevent 44.7% of the modeled six-month BOS-associated costs among additionally detected cases to offset monitoring expenses, whereas universal monitoring would require 104.2% cost reduction and therefore could not break even. Under 30% mitigation and 0.10 assumed annual utility gain, targeted-monitoring NMB was -$54,897 at 5%, +$70,206 at 10%, and +$195,309 at 15% BOS frequency.

**Interpretation:** BOS was associated with a higher post-index institutional-cost trajectory than no extrapulmonary cGVHD and no cGVHD. More frequent PFT surveillance, but not geographic or socioeconomic access, was associated with detection of early declines. Targeted home monitoring became economically favorable only under selected higher-risk assumptions.

## Introduction

Bronchiolitis obliterans syndrome (BOS) is the primary pulmonary manifestation of chronic graft-versus-host disease (cGVHD) after allogeneic hematopoietic cell transplantation (HCT). BOS is diagnosed clinically by persistent airflow obstruction ^1,2^ as defined by National Institutes of Health criteria^3^. BOS occurs in 5.5% of allogeneic HCT recipients overall and 14% of those with cGVHD^3^. A comparable frequency was observed in our institutional parent cohort, in which 156 of 3,170 recipients (4.9%) developed BOS^4^. Detecting BOS in a timely fashion remains challenging. In a cohort of 3,170 HCT recipients, 156 developed BOS, but early pulmonary declines prior to BOS were only detected in 72 patients^4^. Because established airflow obstruction is often only partially reversible, and affected patients may experience morbidity and mortality due to pulmonary impairment^5^ and prolonged immunosuppression, ^6-9^, but efficient detection of BOS is limited by resources allowing for more frequent clinic-based pulmonary function tests (PFTs).

One of the primary challenges in the management of BOS is that BOS is often recognized only after substantial lung-function loss has occurred. An official American Thoracic Society research statement identified late-stage diagnosis and the severity of pulmonary impairment at recognition as central obstacles to improving outcomes^3^. Airflow obstruction often progresses well before symptoms become prominent, highlighting the central role of PFT monitoring^10-12^. The usefulness of PFTs, however, depends on whether PFTs can be performed frequently enough to reliably detect BOS. More broadly, real-world pulmonary surveillance after HCT remains incomplete. In a national claims analysis by our group, only 44% of commercially insured and 36% of Medicare allogeneic HCT recipients underwent any lung-function testing during the first post-transplant year. Among recipients still observable in the third year, these proportions were 31% and 26%, respectively^13^, indicating a major implementation gap for existing guidelines.

Home spirometry may provide a practical way to increase surveillance frequency without requiring repeated laboratory visits. Handheld spirometry correlates with laboratory-based measurements^14^, and home telemonitoring has been shown to be feasible and technically reliable in allogeneic HCT recipients^15^. In a high-risk cGVHD cohort, home spirometry identified lung-function decline before conventional BOS diagnosis in many patients who subsequently developed BOS or suspected BOS^16^. A mixed-methods study by our group also demonstrated the acceptability and usability of home spirometry among HCT recipients^17^. However, the feasibility of home spirometry implementation is inherently tied to the cost of home spirometry monitoring and whether this should be applied to all HCT recipients or only those deemed at high risk for BOS, such as those with extrapulmonary cGVHD.

We therefore examined monthly institutional costs among adult recipients of a first allogeneic HCT with BOS, cGVHD without BOS, or no cGVHD focusing on the date of BOS or non-BOS cGVHD as the index date. We also evaluated the frequency and lead time of antecedent pulmonary decline, factors associated with earlier detection, and the relationship between pulmonary severity at BOS diagnosis and subsequent cost. Because gaps in surveillance may reflect barriers to access, we examined whether proximity to specialized care, socioeconomic vulnerability, or completion of scheduled PFTs was associated with undetected pulmonary impairment among all BOS cases in the parent cohort. Finally, using the observed BOS-associated cost differential as an anchor, we conducted exploratory analyses to identify the conditions under which targeted or universal home spirometry might be economically favorable.

## Methods

### Study Design and Data Sources

We conducted a retrospective case-control study in a cohort of adult recipients with first allogeneic HCT at UT MD Anderson. The parent cohort included 3,170 consecutive adults who underwent their first allogeneic HCT for a primary hematologic malignancy between February 1999 and March 2018^4^. Clinical, demographic, and institutional billing data were linked at the patient level. The study was approved by the MD Anderson Institutional Review Board (PA17-0732) with a waiver of informed consent.

### Study Population

The nested case-control sample included 462 patients: 142 with BOS, 170 with extensive systemic cGVHD without BOS, and 150 without cGVHD. BOS cases were identified using NIH guideline-based diagnostic criteria and were confirmed through expert clinical adjudication^4^. Patients with non-BOS cGVHD and no cGVHD were matched to patients with BOS based upon age (within 10 years), era of transplantation (divided into 5-year quartiles), and time since HCT to the index date (within 1 month). Patients were excluded from the economic analysis if they lacked linked institutional billing or index-date information (n = 104), had undergone one or more prior HCTs (n = 34), had an index date after September 5, 2015 (n = 15), or lacked a valid index-centered cost month (n = 1). The final analysis included 308 patients: 111 with BOS, 132 with cGVHD without BOS, and 65 without cGVHD **(Figure 1)**. All patients in the economic full analysis set had undergone their first allogeneic HCT on or before December 15, 2014; the last eligible index date was August 27, 2015 in order to match normalized billing data.

**Figure 1.**
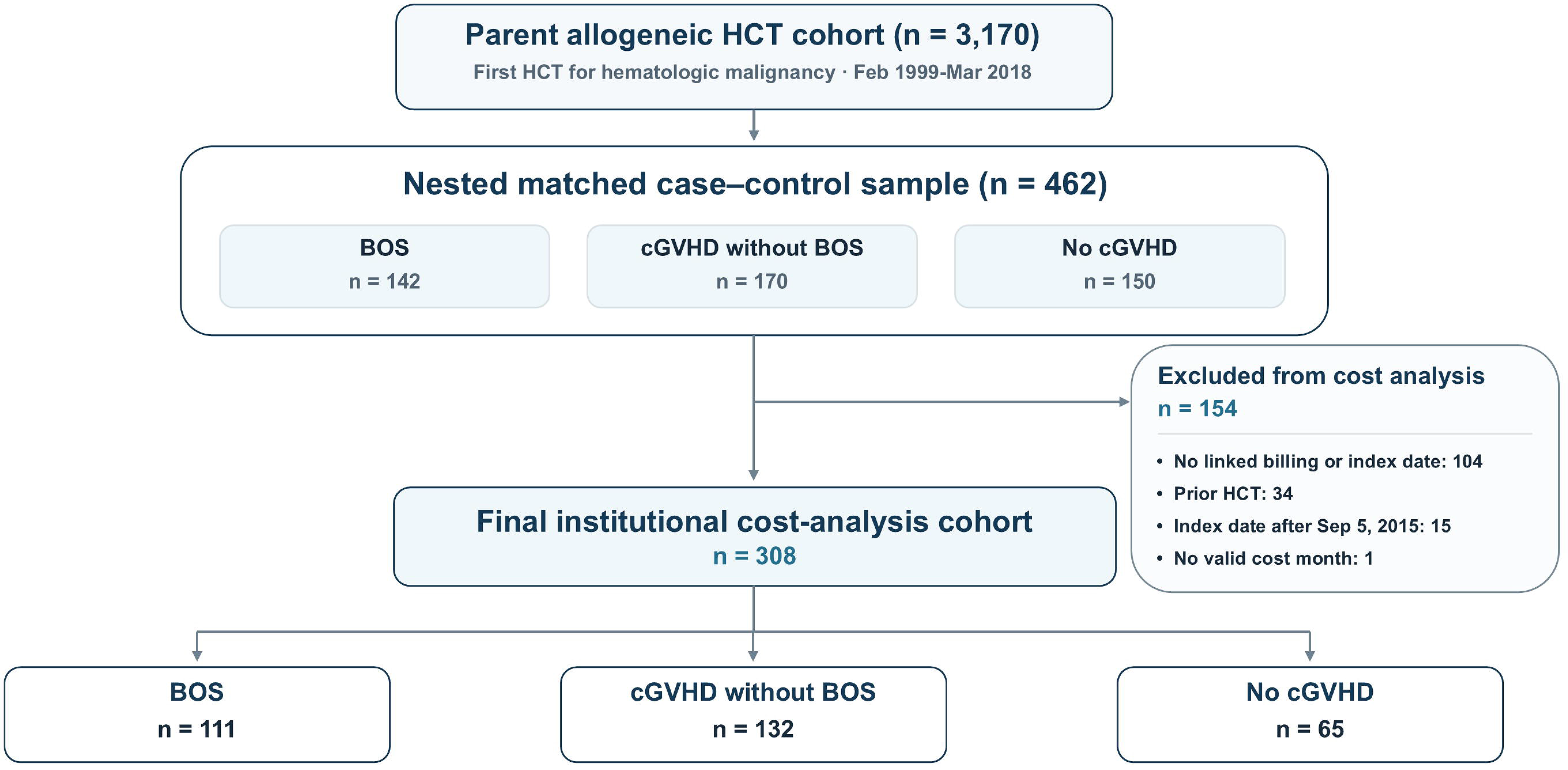
Study Population Selection and Analytic Cohort. The parent cohort comprised 3,170 adults who underwent their first allogeneic hematopoietic cell transplantation (HCT) for hematologic malignancies between February 1999 and March 2018. A nested matched case-control sample included 462 patients: 142 with bronchiolitis obliterans syndrome (BOS), 170 with chronic graft-versus-host disease (cGVHD) without BOS, and 150 without cGVHD. Of these, 154 patients were excluded because of unavailable linked billing or index-date information (n = 104), prior HCT (n = 34), index dates after September 5, 2015 (n = 15), or absence of a valid index-centered cost month (n = 1). The final economic analysis included 308 patients: 111 with BOS, 132 with cGVHD without BOS, and 65 without cGVHD.

### Cost Outcomes and Inflation Adjustment

The primary economic outcome was monthly institutional health-care cost expressed in 2024 US dollars. Nominal costs were converted according to the calendar year in which each service occurred using annual average values from the US Bureau of Labor Statistics Consumer Price Index for All Urban Consumers Medical Care series (CPI-U Medical Care; series CUUR0000SAM; 1982-1984 = 100), where the annual average CPI-U Medical Care value for 2024 was 563.841^18,19^.

### Index-Centered Monthly Observation Windows

Costs were aggregated into 13 calendar-month bins from M-6 through M+6 relative to the index date. M0 represented the calendar month containing the index event. The primary pre-index period was the average of M-6 through M-1, and the primary post-index period was the average of M+1 through M+6. A month without a billing claim was assigned a cost of $0 only when the patient was alive and observable in the linked institutional billing panel. Months occurring before billing observation, after the end of documented follow-up, or after death were treated as unobservable and were not assigned zero cost. The calendar month in which death occurred was retained, but costs were not carried forward after death. Monthly costs were summarized using the number of alive and observable patients, mean, median, interquartile range, and 95% CIs. Mean costs were emphasized for the economic comparisons because they represent population-level resource use. Bootstrap CIs were generated by resampling patients with replacement using 5,000 repetitions.

### PFT Measurements

Baseline pulmonary function was defined as the most recent physiologically plausible PFT performed on or before HCT. Baseline measures included FEV₁ in liters, FEV₁ percent-predicted, FEV₁ z-score, FEV₁/FVC ratio, and FEV₁/FVC z-score. The primary pulmonary-surveillance outcome was whether an antecedent pulmonary decline had been recorded before the adjudicated BOS diagnosis date. Lead time was calculated as the number of days from the recorded antecedent decline to BOS diagnosis. Separately, repeated PFT measurements between HCT and BOS diagnosis were used to identify relative FEV₁ declines of at least 10%, 15%, and 20% from the baseline FEV₁. For each threshold, the first qualifying PFT before BOS was identified and its lead time to BOS diagnosis was calculated. Thresholds were nested; therefore, a patient reaching a 20% decline also satisfied the 10% and 15% thresholds. Surveillance intensity was defined as the number of PFTs performed before BOS divided by the interval from HCT to BOS in years.

### Statistical Analysis

All analyses were performed using R version 4.5.2. Continuous baseline variables were summarized using means, medians and interquartile ranges and compared descriptively using Kruskal-Wallis tests. Categorical variables were summarized using counts and percentages and compared using Fisher exact tests.

#### Longitudinal cost analysis

Monthly cost was analyzed using a Gaussian identity-link generalized estimating equation (GEE) with patient as the repeated cluster, an exchangeable working correlation structure, and robust sandwich standard errors. Month was modeled as a 13-level categorical variable from M-6 through M+6. The model included clinical group, categorical month, and the group-by-month interaction.

The identity link preserved interpretation in absolute 2024 US dollars. Marginal mean costs and covariance-based linear contrasts were estimated from the complete GEE model. Prespecified contrasts included within-group adjacent-month changes, pairwise between-group comparisons within each relative month, within-group differences between average post-index and pre-index monthly costs, and difference-in-differences comparisons between the pre/post changes of the three groups. A positive BOS-vs-control difference-in-differences estimate indicated that the BOS cost trajectory was more costly, or less favorable, than the corresponding control trajectory.

#### Antecedent-decline analysis

Detection proportions and FEV₁ threshold-crossing proportions were reported with exact binomial 95% CIs. Lead times were summarized using medians. Associations between pulmonary surveillance and antecedent-decline detection were evaluated using ordinary logistic regression. The four primary surveillance exposures were PFTs per patient-year and completion of the 3-, 6-, and 12-month PFTs. Scheduled-PFT models were restricted to patients eligible at the corresponding landmark. Odds ratios were reported with profile-likelihood 95% CIs.

#### Undetected-impairment analysis

Undetected pulmonary impairment was defined as BOS diagnosed without a previously identified antecedent pulmonary decline, that is, no prior PFT showing a >10% decline in FEV₁. Geographic exposures were residence within 10, 15, or 30 miles of a National Cancer Institute (NCI)-designated cancer center or a pulmonologist, and drive time and distance to the nearest NCI-designated center. Socioeconomic exposures were the Centers for Disease Control and Prevention Social Vulnerability Index (SVI) percentile rank, income above the 2023 US Department of Labor median of $48,060, and college or university education. Surveillance exposures were completion of the day-100, 6-month, and 12-month PFTs. Associations were estimated using univariable logistic regression with Wald 95% CIs.

#### Pulmonary severity and subsequent cost

Among BOS cases, mean monthly cost during M+1 through M+6 was analyzed using Ordinary Least Squares (OLS) regression with HC3 robust standard errors. Models evaluated documented antecedent decline, FEV₁ at BOS diagnosis, magnitude of serial FEV₁ decline, and FEV₁ z-score.

### Exploratory Home-Spirometry Economic Analysis

We performed an exploratory deterministic six-month cost-utility and net monetary benefit (NMB) analysis of supplemental home spirometry in a hypothetical cohort of 400 HCT recipients. The primary economic alert was a ≥10% relative FEV₁ decline. Current clinic surveillance identified this threshold before BOS in 35% of local BOS cases. Weekly home spirometry was assigned an external observed detection benchmark of 75%^16^, yielding an incremental detection assumption of 40%; the external estimate was not treated as a head-to-head comparison.

Two strategies were evaluated: targeted monitoring of 30% of recipients, assumed to contain 70% of future BOS cases, and universal monitoring of all recipients. Monitoring cost was $250 per monitored patient-month for six months, corresponding to $180,000 for targeted and $600,000 for universal monitoring. Assumed BOS cohort frequencies were 2%, 3%, 5%, 10%, and 15%. The six-month BOS-associated cost anchor was $72,313, derived from the final unadjusted BOS-versus-no-cGVHD DiD.

For each strategy and BOS frequency, we calculated additional early detections, the proportion of the BOS-associated cost burden that would need to be mitigated for cost break-even, and the maximum monthly monitoring price compatible with break-even. For exploratory cost-utility analyses, cost mitigation after additional early detection was varied across 10%, 30%, and 50%, and assumed annual utility gain per effectively mitigated BOS case was varied across 0, 0.05, 0.10, and 0.20. QALY gains were accrued over the six-month horizon as effective cases x annual utility gain x 0.5. Incremental cost was monitoring cost minus avoided BOS-associated cost. NMB was calculated as ($50,000 x QALYs gained) minus incremental cost; NMB >$0 was classified as positive NMB.

## Results

### Baseline Characteristics

The overall median age at HCT was 49 years (IQR, 37-57), and 180 patients (58%) were male **(Table 1)**. AML or MDS was the most common malignancy, affecting 148 patients (48%). Age, sex, malignancy, HCT era, and donor type did not differ substantially among the three groups. Baseline pulmonary function differed across groups **(Table 1)**. Patients who developed BOS had lower baseline FEV₁ z-scores (median, -0.43 vs -0.09 in cGVHD and -0.14 in no cGVHD; p = 0.009) and FEV₁/FVC z-scores (median, -0.32 vs 0.29 and 0.11, respectively; p < 0.001).

**Table 1.** Baseline Characteristics of the Economic Full Analysis Set by Clinical Group.

| **Characteristic** | **Overall<br>(N = 308)** | **No cGVHD<br>(n = 65)** | **cGVHD<br>Without BOS<br>(n = 132)** | **BOS<br>(n = 111)** | **p Value** |
|---|---|---|---|---|---|
| Age at HCT, y | 49 (37-57) | 50 (39-59) | 47 (38-56) | 50 (35-57) | 0.5 |
| Sex |  |  |  |  | 0.5 |
| Female | 128 (42) | 31 (48) | 53 (40) | 44 (40) |  |
| Male | 180 (58) | 34 (52) | 79 (60) | 67 (60) |  |
| Malignancy |  |  |  |  | 0.4 |
| AML/MDS | 148 (48) | 28 (43) | 60 (45) | 60 (54) |  |
| Lymphoma | 51 (17) | 16 (25) | 18 (14) | 17 (15) |  |
| CML/MPD | 30 (9.7) | 3 (4.6) | 14 (11) | 13 (12) |  |
| ALL | 26 (8.4) | 7 (11) | 13 (9.8) | 6 (5.4) |  |
| CLL | 23 (7.5) | 5 (7.7) | 13 (9.8) | 5 (4.5) |  |
| Myeloma/plasma cell | 8 (2.6) | 1 (1.5) | 5 (3.8) | 2 (1.8) |  |
| Other | 22 (7.1) | 5 (7.7) | 9 (6.8) | 8 (7.2) |  |
| Race and ethnicity |  |  |  |  | 0.038 |
| White | 241 (78) | 49 (75) | 103 (78) | 89 (80) |  |
| Black | 15 (4.9) | 6 (9.2) | 5 (3.8) | 4 (3.6) |  |
| Hispanic | 22 (7.1) | 0 | 10 (7.6) | 12 (11) |  |
| Asian | 5 (1.6) | 1 (1.5) | 3 (2.3) | 1 (0.9) |  |
| Other | 22 (7.1) | 8 (12) | 9 (6.8) | 5 (4.5) |  |
| Unknown | 3 (1.0) | 1 (1.5) | 2 (1.5) | 0 |  |
| HCT era |  |  |  |  | 0.4 |
| 1999-2004 | 79 (26) | 11 (17) | 34 (26) | 34 (31) |  |
| 2005-2009 | 88 (29) | 21 (32) | 36 (27) | 31 (28) |  |
| 2010-2014 | 141 (46) | 33 (51) | 62 (47) | 46 (41) |  |
| Donor type |  |  |  |  | 0.8 |
| MRD | 191 (62) | 39 (60) | 81 (61) | 71 (64) |  |
| MUD | 117 (38) | 26 (40) | 51 (39) | 40 (36) |  |
| Baseline pulmonary function |  |  |  |  |  |
| FEV<sub>1</sub>, L | 3.07 (2.61-3.77) | 2.97 (2.60-3.70) | 3.26 (2.74-3.89) | 3.00 (2.51-3.66) | 0.03 |
| FEV<sub>1</sub>, % predicted | 96 (88-107) | 98 (91-107) | 99 (90-109) | 94 (81-103) | 0.009 |
| FEV<sub>1</sub> z score | -0.27 (-0.84 to 0.45) | -0.14 (-0.61 to 0.50) | -0.09 (-0.71 to 0.58) | -0.43 (-1.31 to 0.20) | 0.009 |
| FEV<sub>1</sub>/FVC | 0.81 (0.76-0.85) | 0.81 (0.78-0.84) | 0.83 (0.79-0.86) | 0.79 (0.74-0.84) | <0.001 |
| FEV<sub>1</sub>/FVC z score | 0.02 (-0.74 to 0.53) | 0.11 (-0.44 to 0.43) | 0.29 (-0.33 to 0.81) | -0.32 (-0.98 to 0.24) | <0.001 |
| Time from HCT to index, mo | 13 (7-23) | 18 (8-24) | 11 (6-18) | 17 (8-27) | <0.001 |

ALL = acute lymphoblastic leukemia; AML = acute myeloid leukemia; BOS = bronchiolitis obliterans syndrome; cGVHD = chronic graft-vs-host disease; CLL = chronic lymphocytic leukemia; CML = chronic myeloid leukemia; HCT = hematopoietic cell transplantation; MDS = myelodysplastic syndrome; MPD = myeloproliferative disorder; MRD = matched related donor; MUD = matched unrelated donor; PFT = pulmonary function test.
Data are No. (%) or median (IQR). p-values are from Kruskal-Wallis tests for continuous variables and Fisher exact tests for categorical variables.

### Monthly Cost Patterns

At M+1, raw mean monthly cost was $13,848 in the no-cGVHD group, $39,337 in the cGVHD group, and $31,796 in the BOS group. By M+5, corresponding raw mean costs were $7,764, $18,695, and $21,548. During M+1 through M+6, 5 patients without cGVHD, 16 with cGVHD, and 18 with BOS died, reducing the number of observable patient-months over time.

### Primary Six-Month Cost Analysis

In the longitudinal GEE, mean monthly institutional cost declined significantly after index in patients without cGVHD (-$7,582; 95% CI, -$13,378 to -$1,786; p =0.01), whereas the changes in patients with cGVHD or BOS were not statistically significant **(Table 2)**. The BOS pre/post trajectory was $12,052/month less favorable than the no-cGVHD trajectory (95% CI, $2,700-$21,404; p=0.01), equivalent to a six-month increased burden of $72,313. BOS was also $10,238/month less favorable than cGVHD before multiplicity correction (95% CI, $330-$20,146; p=0.04); the six-month increase in cost was $61,426. The cGVHD-versus-no-cGVHD DiD was $1,814/month (95% CI, -$7,012 to $10,641; p=0.68). The within-group cost changes and between-group DiD estimates are summarized in **Figure 2**.

**Figure 2.**
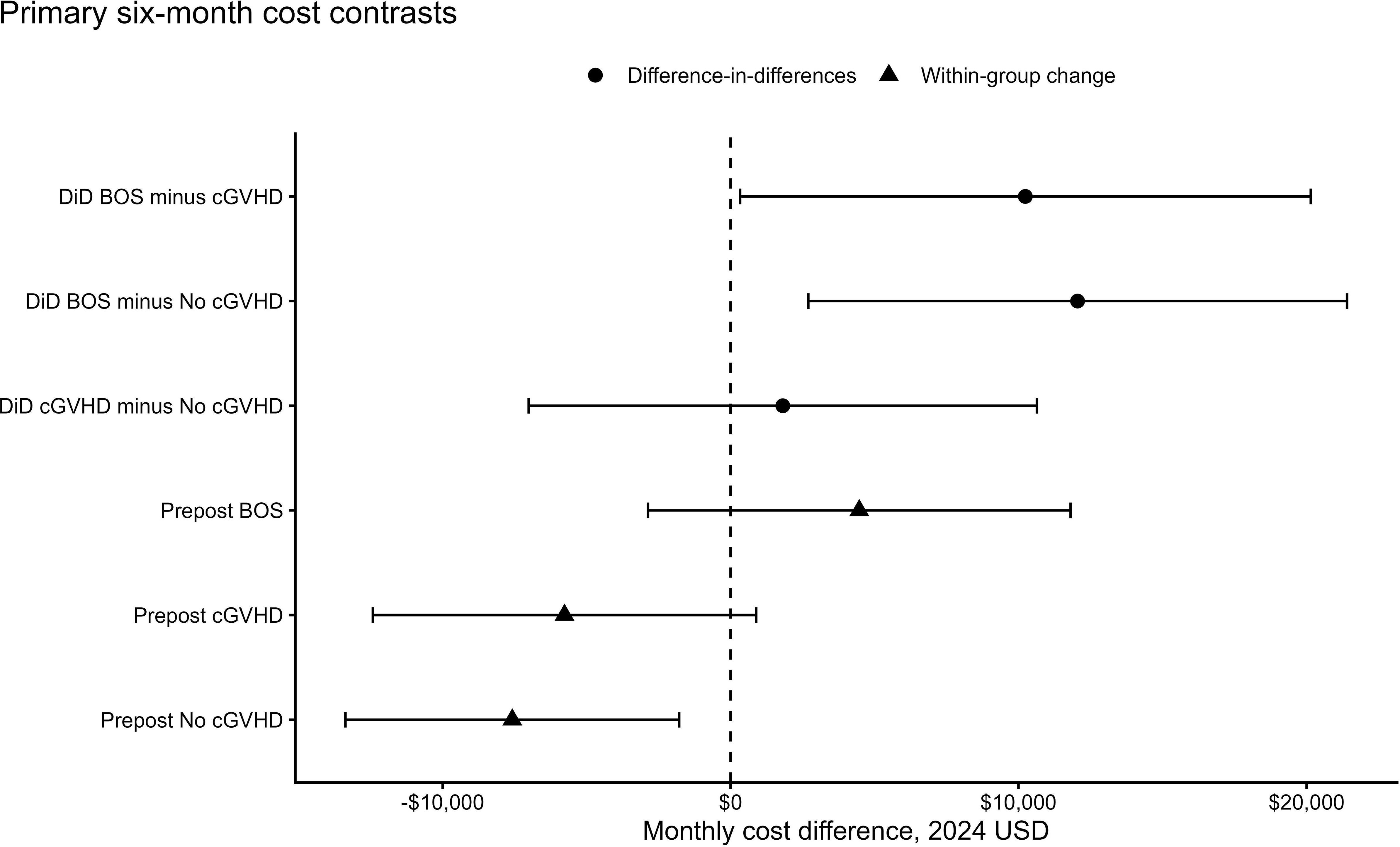
Six-Month Changes in Institutional Costs and Difference-in-Differences Comparisons Following Allogeneic Hematopoietic Cell Transplantation. Forest plot showing within-group changes in mean monthly institutional costs and between-group difference-in-differences (DiD) estimates among 308 allogeneic hematopoietic cell transplantation recipients, including 111 with bronchiolitis obliterans syndrome (BOS), 132 with chronic graft-versus-host disease (cGVHD) without BOS, and 65 without cGVHD. Costs were compared between the six-month pre-index (M-6 to M-1) and post-index (M+1 to M+6) periods using generalized estimating equations and are expressed in inflation-adjusted 2024 US dollars. Circles represent between-group DiD estimates, triangles indicate within-group pre-to-post changes, and horizontal lines represent 95% confidence intervals (CIs). Positive values indicate an increase in monthly costs or a less favorable cost trajectory relative to the comparison group. The dashed vertical line represents no difference ($0).

**Table 2.** Six-Month Pre/Post Change in Mean Monthly Institutional Cost and Difference-in-Differences by Clinical Group.

| **Comparison** | **Monthly Estimate, \$<br>(95% CI)** | **Six-Month<br>Estimate, \$** | **p Value** | **Adjusted<br>p Value** |
|---|---|---|---|---|
| **Within-group change (post-index minus pre-index)** |  |  |  |  |
| No cGVHD (n = 65) | -7,582 (-13,378 to -1,786) | -45,491 | 0.01 | 0.03 |
| cGVHD without BOS (n = 132) | -5,768 (-12,424 to 889) | -34,605 | 0.09 | 0.18 |
| BOS (n = 111) | 4,470 (-2,869 to 11,809) | 26,821 | 0.23 | 0.23 |
| **Difference-in-differences** |  |  |  |  |
| BOS vs no cGVHD | 12,052 (2,700 to 21,404) | 72,313 | 0.01 | 0.03 |
| BOS vs cGVHD without BOS | 10,238 (330 to 20,146) | 61,426 | 0.04 | 0.09 |
| cGVHD without BOS vs no cGVHD | 1,814 (-7,012 to 10,641) | 10,886 | 0.69 | 0.69 |

BOS = bronchiolitis obliterans syndrome; cGVHD = chronic graft-vs-host disease; CPI-U = Consumer Price Index for All Urban Consumers; GEE = generalized estimating equation; M = month relative to the index date.
Costs are monthly institutional costs in 2024 US dollars (CPI-U Medical Care). Estimates are linear contrasts from a Gaussian identity-link GEE that included clinical group, month as a 13-level categorical variable (M−6 through M+6), and the group-by-month interaction, with an exchangeable working correlation and robust sandwich standard errors.
The pre-index and post-index periods were the means of M−6 through M−1 and M+1 through M+6, respectively.
Within-group change is post-index minus pre-index mean monthly cost. Difference-in-differences is the difference between the within-group changes of the 2 groups compared; positive values indicate a less favorable cost trajectory in the first group listed. Six-month estimates are the monthly estimates multiplied by 6.
Adjusted p values are Holm-corrected separately for the 3 within-group comparisons and the 3 difference-in-differences comparisons.

### Pulmonary Surveillance and Antecedent Decline

A clinically antecedent decline was detected before BOS in 40 of 108 patients, corresponding to 37% (95% CI, 28%-47%). Among patients with detected decline, the median lead time before BOS diagnosis was 268 days. PFT completion improved in modern HCT eras. Completion of the 3-month PFT increased from 1 of 32 eligible patients (3.1%) in 1999-2004 to 28 of 41 (68.3%) in 2010-2014. Completion of the 6-month PFT increased from 2 of 30 (6.7%) to 15 of 38 (39.5%), and completion of the 12-month PFT increased from 4 of 24 (16.7%) to 14 of 29 (48.3%). Each additional PFT per patient-year was associated with more than two-fold higher odds of detecting antecedent decline (OR, 2.4; 95% CI, 1.4-4.0; p=0.001).

### FEV₁ Threshold Detection

Before BOS diagnosis, 38 of 108 patients (35%; 95% CI, 26.2%-45.0%) had a 10% relative FEV₁ decline in the PFT prior to BOS diagnosis. The median interval between threshold crossing and BOS diagnosis was 267.5 days. A 15% decline was identified in 34 of 108 patients (31.5%; 95% CI, 22.9%-41.1%), with a median lead time of 217 days. A 20% decline was identified in 30 of 108 patients (27.8%; 95% CI, 19.6%-37.2%), with a median lead time of 190 days.

### Correlates of Undetected Pulmonary Impairment

Undetected antecedent impairment was not significantly associated with residence within 30 miles of an NCI-designated center (OR, 0.88; 95% CI, 0.42-1.88; p=0.75) or a pulmonologist (OR, 0.66; 95% CI, 0.21-2.06; p=0.47), drive time (OR, 0.95 per hour; 95% CI, 0.75-1.19; p=0.63), or driving distance (OR, 0.95 per 100 km; 95% CI, 0.77-1.17; p=0.63). Results were similar at 10- and 15-mile thresholds.

The median SVI percentile rank was 0.43 (IQR, 0.23-0.68); 53.7% of patients had income above the national median, and 46.9% had a college or university education. Undetected impairment was not significantly associated with SVI (OR, 1.02 per 10 percentiles; 95% CI, 0.91-1.15; p=0.71), income above the national median (OR, 1.64; 95% CI, 0.85-3.16; p=0.14), or college or university education (OR, 0.80; 95% CI, 0.42-1.54; p=0.50).

Undetected antecedent impairment became less frequent across eras, from 74.4% in 1999-2004 to 54.3% in 2005-2009, 53.4% in 2010-2014, and 13.3% thereafter (p<0.001). Completion of scheduled PFTs was associated with lower unadjusted odds of undetected antecedent impairment at day 100 (OR, 0.40; 95% CI, 0.20-0.81; p=0.01), 6 months (OR, 0.31; 95% CI, 0.16-0.62; p<0.001), and 12 months (OR, 0.41; 95% CI, 0.21-0.80; p=0.009) **(Figure 3)**, with similar estimates after restriction to patients without BOS at each landmark (ORs, 0.46, 0.33, and 0.44, respectively). After adjustment for era, the association persisted for the 6-month PFT (OR, 0.38; 95% CI, 0.18-0.79; p=0.01) but was attenuated for the day-100 (OR, 0.69; 95% CI, 0.26-1.84; p=0.46) and 12-month PFTs (OR, 0.58; 95% CI, 0.28-1.19; p=0.14). These findings were directionally consistent with the association between clinic PFT frequency and antecedent-decline detection described above.

**Figure 3.**
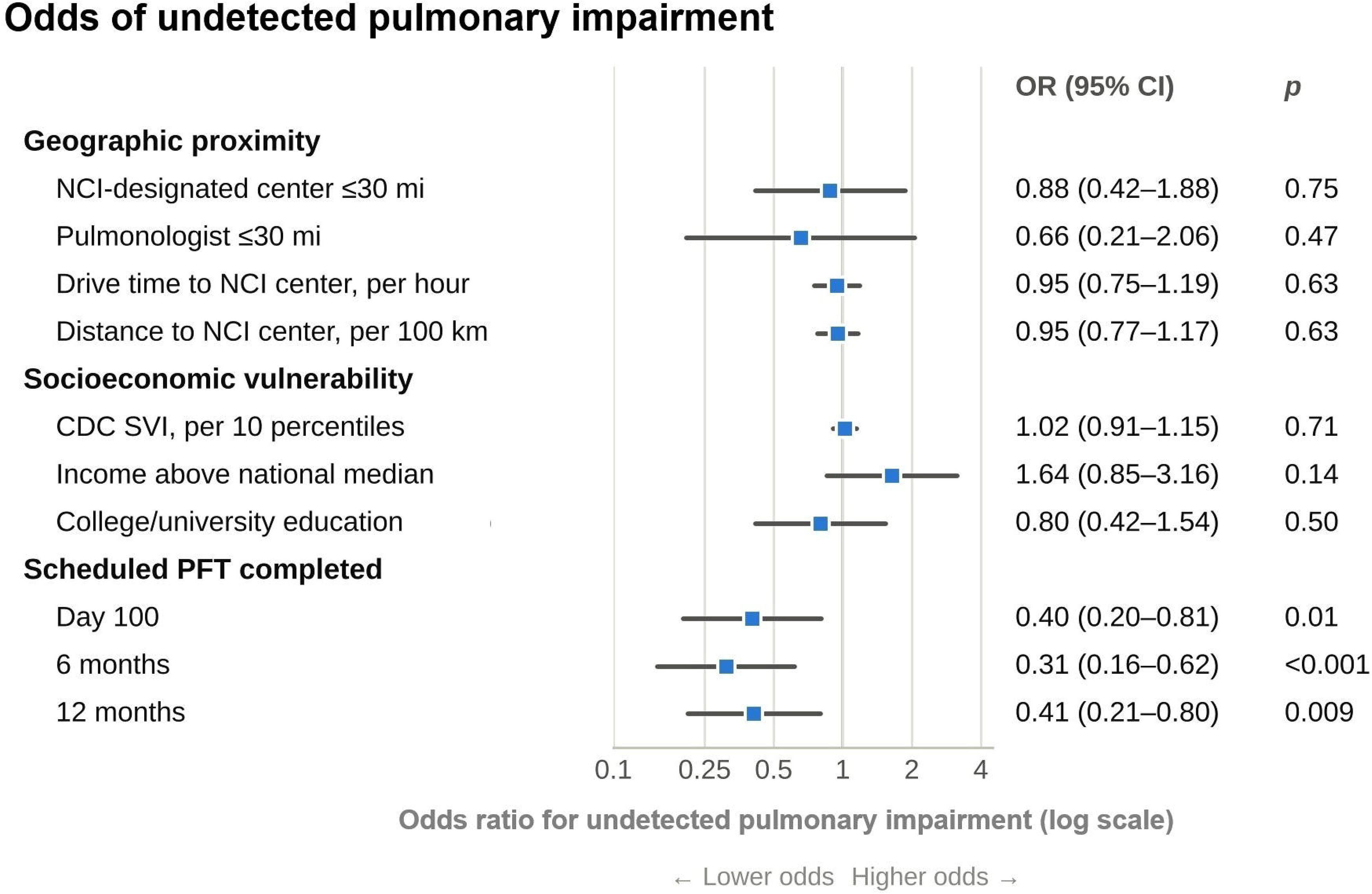
Associations of Geographic Accessibility, Socioeconomic Vulnerability, and Pulmonary Surveillance With Undetected Pulmonary Impairment. Forest plot showing associations between geographic accessibility, socioeconomic characteristics, scheduled pulmonary function test (PFT) completion, and undetected antecedent pulmonary impairment among patients diagnosed with bronchiolitis obliterans syndrome (BOS) following allogeneic hematopoietic cell transplantation. Undetected impairment was defined as the absence of a previously documented PFT demonstrating a greater than 10% decline in forced expiratory volume in 1 second (FEV₁) before BOS diagnosis. Odds ratios (ORs) and 95% confidence intervals (CIs) were estimated using separate univariable logistic regression models. Geographic exposures included proximity to a National Cancer Institute (NCI)-designated cancer center or pulmonologist, driving time, and travel distance. Socioeconomic characteristics included the Centers for Disease Control and Prevention Social Vulnerability Index (SVI), income, and educational attainment.

### Pulmonary Severity and Subsequent Cost

In exploratory BOS-only analyses, greater pulmonary impairment at BOS diagnosis was associated with higher subsequent cost. Each 10-percentage-point lower FEV₁ percent predicted at the time of BOS diagnosis was associated with $6,467 higher subsequent mean monthly cost (95% CI, $445–$12,490; p = 0.03), whereas each additional 10% relative FEV₁ decline from baseline to BOS diagnosis was associated with $5,011 higher monthly cost (95% CI, $1,178-$8,844; p = 0.01); and each 1-SD lower FEV₁ z score with +$13,115/month additional cost (95% CI, $4,807-$21,424; p=0.002). Patients with a documented antecedent decline had a mean monthly subsequent cost $13,027 lower than those without (95% CI, -$27,718 to $1,664; p=0.08).

### Exploratory Home-Spirometry Scenarios

We modeled two home-spirometry strategies in a hypothetical cohort of 400 HCT recipients. Targeted monitoring included 120 patients (30%) considered at higher risk for BOS, assumed to account for 70% of future BOS cases, whereas universal monitoring included all 400 recipients. At $250 per patient per month, six months of monitoring cost $180,000 for the targeted strategy and $600,000 for universal monitoring. Conventional clinic surveillance identified a ≥10% FEV₁ decline before BOS diagnosis in 35.2% of cases, compared with an external 75% detection benchmark using weekly home spirometry^16^. We therefore assumed that home monitoring would increase early detection by approximately 40 percentage points. At an assumed BOS frequency of 5% (20 of 400 recipients), targeted monitoring was projected to identify antecedent pulmonary decline in 5.57 additional future BOS cases, compared with 7.96 additional cases under universal monitoring. To offset monitoring expenses, targeted monitoring would need to prevent 44.7% of the modeled six-month BOS-associated costs for each additionally detected case, whereas universal monitoring would require a 104.2% reduction and could not break even. Results across other assumed BOS frequencies are presented in **Table 3**.

**Table 3.**
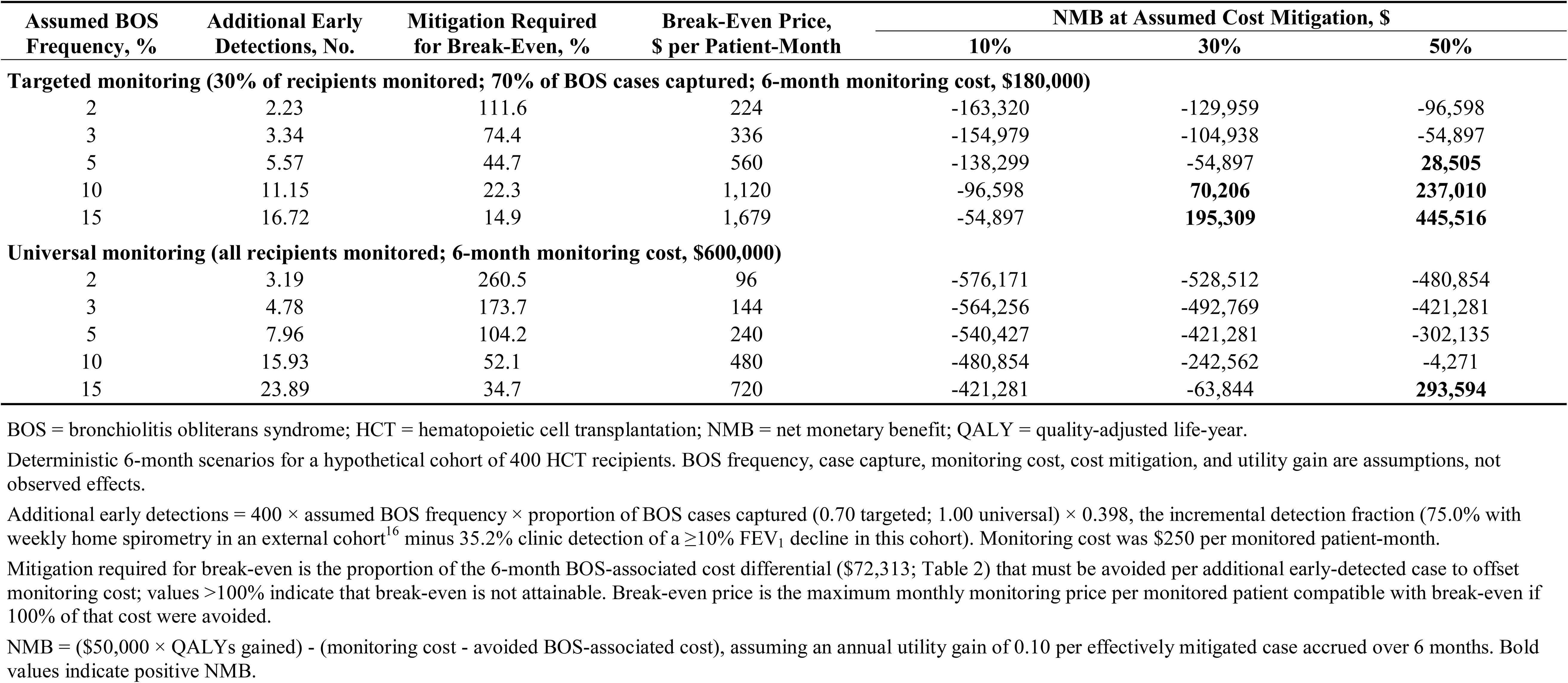
Exploratory Six-Month Break-Even and Net Monetary Benefit of Targeted and Universal Home Spirometry by Assumed BOS Frequency.

| Assumed BOS Frequency, % | Additional Early Detections, No. | Mitigation Required for Break-Even, % | Break-Even Price, \$ per Patient-Month | NMB at Assumed Cost Mitigation, \$ |  |  |
|---|---|---|---|---|---|---|
|  |  |  |  | 10% | 30% | 50% |
| **Targeted monitoring (30% of recipients monitored; 70% of BOS cases captured; 6-month monitoring cost, \$180,000)** |  |  |  |  |  |  |
| 2 | 2.23 | 111.6 | 224 | -163,320 | -129,959 | -96,598 |
| 3 | 3.34 | 74.4 | 336 | -154,979 | -104,938 | -54,897 |
| 5 | 5.57 | 44.7 | 560 | -138,299 | -54,897 | **28,505** |
| 10 | 11.15 | 22.3 | 1,120 | -96,598 | **70,206** | **237,010** |
| 15 | 16.72 | 14.9 | 1,679 | -54,897 | **195,309** | **445,516** |
| **Universal monitoring (all recipients monitored; 6-month monitoring cost, \$600,000)** |  |  |  |  |  |  |
| 2 | 3.19 | 260.5 | 96 | -576,171 | -528,512 | -480,854 |
| 3 | 4.78 | 173.7 | 144 | -564,256 | -492,769 | -421,281 |
| 5 | 7.96 | 104.2 | 240 | -540,427 | -421,281 | -302,135 |
| 10 | 15.93 | 52.1 | 480 | -480,854 | -242,562 | -4,271 |
| 15 | 23.89 | 34.7 | 720 | -421,281 | -63,844 | **293,594** |

BOS = bronchiolitis obliterans syndrome; HCT = hematopoietic cell transplantation; NMB = net monetary benefit; QALY = quality-adjusted life-year.
Deterministic 6-month scenarios for a hypothetical cohort of 400 HCT recipients. BOS frequency, case capture, monitoring cost, cost mitigation, and utility gain are assumptions, not observed effects.
Additional early detections = 400 × assumed BOS frequency × proportion of BOS cases captured (0.70 targeted; 1.00 universal) × 0.398, the incremental detection fraction (75.0% with weekly home spirometry in an external cohort<sup>16</sup> minus 35.2% clinic detection of a ≥10% FEV<sub>1</sub> decline in this cohort). Monitoring cost was \$250 per monitored patient-month.
Mitigation required for break-even is the proportion of the 6-month BOS-associated cost differential (\$72,313; Table 2) that must be avoided per additional early-detected case to offset monitoring cost; values >100% indicate that break-even is not attainable. Break-even price is the maximum monthly monitoring price per monitored patient compatible with break-even if 100% of that cost were avoided.
NMB = (\$50,000 × QALYs gained) - (monitoring cost - avoided BOS-associated cost), assuming an annual utility gain of 0.10 per effectively mitigated case accrued over 6 months. Bold values indicate positive NMB.

NMB was strongly dependent on BOS frequency and assumed mitigation. Under the illustrative scenario of 30% cost mitigation, 0.10 assumed annual utility gain, and a $50,000/QALY willingness-to-pay threshold, targeted monitoring yielded NMBs of -$54,897 at 5% BOS frequency, +$70,206 at 10%, and +$195,309 at 15%. Under the same assumptions, universal monitoring was not cost effective at 15% BOS frequency (NMB, -$63,844). At a 5% BOS frequency, targeted monitoring achieved positive NMB only when assumed cost mitigation reached 50%; NMB ranged from +$21,538 with no utility gain to +$35,473 with an annual utility gain of 0.20. Universal monitoring remained negative across all modeled mitigation and utility assumptions at 5% BOS frequency. These findings represent projections based on assumed BOS risk, monitoring costs, earlier detection, and potential treatment benefits rather than cost savings directly observed in patients.

## Discussion

In this study of adult first allogeneic HCT recipients, three findings characterize the economic impact of BOS. First, the institutional-cost trajectory after BOS diagnosis was higher than in matched patients with systemic cGVHD and those with no cGVHD. Compared to patients without cGVHD, the BOS trajectory was $12,052/month less favorable from the pre-index to the post-index period, corresponding to a six-month differential burden of $72,313. Second, among patients with BOS, greater pulmonary impairment was associated with higher cost trajectories; lower FEV₁ z-score at diagnosis and greater decline from an individual patient’s baseline were each associated with greater mean monthly costs during the following six months. Third, pulmonary decline was often detectable months before BOS diagnosis, and more frequent clinic PFT surveillance was associated with a greater likelihood of recognizing that decline. Together, these findings suggest that the value of pulmonary surveillance depends not simply on earlier detection but on whether that lead time can be converted into clinical action that preserves lung function and reduces subsequent disease burden.

The economic impact of BOS was most apparent in the divergence of cost trajectories surrounding the index date. Mean monthly cost declined significantly after index among patients without cGVHD, whereas no significant pre-to-post change occurred in the BOS group. The resulting BOS-versus-no-cGVHD difference-in-differences was $12,052 per month, or $72,313 over six months. The BOS trajectory was also $10,238 per month less favorable than that of patients with systemic extrapulmonary cGVHD. The absence of a significant cGVHD-versus-no-cGVHD difference-in-differences should not be interpreted as evidence that cGVHD itself carries little economic burden. Patients with cGVHD continued to incur substantial absolute costs, and the comparator index dates were designed to align observation periods rather than capture the cumulative economic burden of cGVHD. Prior studies have demonstrated substantial treatment costs and health-care utilization associated with cGVHD and pulmonary complications after HCT^20-22^. The present analysis shows that the post-index decline in costs observed in the comparator groups was not seen after BOS diagnosis.

The association between pulmonary impairment and subsequent cost provides an important link between the clinical and economic findings. Among patients with BOS, each additional 10% relative FEV₁ decline from baseline was associated with approximately $5,011 higher mean monthly costs during the subsequent six months. Each 10-percentage-point decrease in FEV₁ percent predicted at BOS was associated with $6,467 higher monthly costs, while each 1-SD decrease in FEV₁ z score was associated with $13,115 higher monthly costs. Prior work has linked more advanced pulmonary impairment in BOS with worse quality of life and mortality^5,9^. Our findings suggest that greater impairment is also associated with greater economic burden. Our findings show that preservation of lung function is not only important from the perspective of minimizing harm to individuals, but can also mitigate downstream costs, highlighting the importance of strategies which facilitate earlier recognition from both patient and payor perspectives.

A clinically documented antecedent pulmonary decline was identified before BOS in 37% of cases with conventional guideline-concordant PFTs and preceded BOS diagnosis by a median of approximately nine months. Serial FEV₁ measurements showed a similar pattern. A relative decline of at least 10% was identified before BOS diagnosis in 35% of patients, with a median lead time of 268 days, while 15% and 20% declines were identified in 32% and 28%, respectively. Surveillance intensity was closely associated with whether this window was recognized. Each additional clinic PFT per patient-year was associated with more than twice the odds of detecting antecedent decline (OR, 2.38; 95% CI, 1.41-4.02; p=0.001). However, current guidelines do not dictate whether these declines should be intervened upon, and as a result in this historical cohort, all patients eventually developed BOS^2^. The contemporary treatment landscape may be more favorable for interventional strategies. Studies of ruxolitinib and belumosudil have reported more favorable pulmonary responses in patients with less advanced lung disease, raising the possibility that treatment may be more effective before severe impairment develops^23,24^. Patients with a documented antecedent decline had an estimated mean monthly subsequent cost $13,027 lower than those without a documented decline, but the association did not meet statistical significance. This observation is sobering but also highlights the potential of intervention and the implementation of prognostic tools such as imaging which can distinguish patients with early BOS from false positives^25^.

Among all BOS cases, undetected impairment was not significantly associated with proximity to specialized care or with socioeconomic vulnerability. Missed recognition therefore appeared more closely related to surveillance patterns rather than barriers to access, though this may be biased by our single-institution design, since patients who are seen at our institution may not be representative of all patients who are evaluated for HCT. These findings should focus implementation efforts on improving surveillance efficacy and efficiency.

The exploratory home-spirometry analysis identifies a potential opportunity to translate earlier detection into economic value. Clinic surveillance identified a ≥10% FEV₁ decline before BOS in 35% of cases, compared with a 75% detection benchmark from 9 of 12 patients monitored with weekly home spirometry in an external cohort^16^. Using this 40% difference as an incremental detection assumption, targeted monitoring was consistently more economically favorable than universal monitoring across all modeled BOS frequencies. At an assumed BOS frequency of 5%, targeted monitoring would need to mitigate 45% of the modeled BOS-associated cost burden to break even, with progressively less mitigation required as BOS risk increased. These findings suggest that concentrating home spirometry among patients at higher risk for BOS, such as those with recent respiratory viral infection^26^ or extrapulmonary cGVHD^4^, may improve the economic value of surveillance and provide a viable strategy for capturing pulmonary decline during a potentially actionable window.

The economic implications of implementing targeted home spirometry should factor in the estimated lifetime direct medical costs of $942,373 to $1,247,917 per allogeneic HCT recipient, with 37% to 53% attributed to cGVHD treatment^20^. French administrative data similarly demonstrated greater health-care utilization and direct costs among patients with GVHD than among recipients without GVHD^21^. Our prior work based upon claims analyses found that noninfectious pulmonary complications incurred mean total costs that were $155,470 higher during the first post-transplant year than those of matched recipients without these complications^22^. Our current work extends those findings because of the universal availability of clinical data allowing for accurate event ascertainment and adjustment for appropriate clinical factors. The increased economic burden imposed by BOS justify efforts to improve BOS surveillance and mitigate further harms to a vulnerable population.

This study has several strengths. Clinical, PFT, vital status, and institutional billing data were linked at the patient level, allowing surveillance, pulmonary deterioration, and cost to be evaluated within the same individuals. BOS cases were confirmed through expert clinical adjudication rather than administrative diagnosis codes. Including both cGVHD without BOS and no-cGVHD comparator groups allowed the BOS-associated trajectory to be distinguished from broader patterns of resource utilization after transplantation. Costs were obtained at each encounter. Several limitations qualify these findings. The retrospective design cannot establish causal relationships among surveillance, pulmonary decline, treatment, and subsequent cost. Institutional billing reflects care captured within a single health system and does not represent total health-care or societal costs; costs incurred outside our institution were not captured. The cohort also reflects an earlier transplantation era, before contemporary cGVHD therapies were widely available. Finally, the economic analyses should be considered exploratory, and the economic scenarios depend on assumptions regarding BOS frequency, monitoring cost, incremental detection, mitigation of downstream burden, and health utility.

## Conclusion

BOS was associated with higher institutional-cost trajectories than other HCT recipients. Among patients with BOS, greater pulmonary impairment at diagnosis was associated with higher subsequent cost. Pulmonary decline was often detectable months before diagnosis but not intervened upon due to the lack of clarity on best practices for early impairment. Increased clinic-based PFTs were associated with a higher odds of antecedent impairment prior to BOS. Exploratory modeling suggested that targeted home spirometry may be economically favorable in higher-risk populations, but only if earlier detection leads to intervention that preserves lung function, a premise that prospective studies should test. Our data justify efforts to implement home spirometry in high-risk populations to identify and intervene upon early pulmonary impairment.

## Data Availability

All data produced in the present study are available upon reasonable request to the authors

## Author Contributions

AMA and AS contributed equally to this work. AMA, AS, and ASh contributed to study conception, design, and methodology. AMA and AS performed data analysis, interpreted the results, prepared the figures and tables, and drafted the manuscript. RP, HEL, CL, HS, RR, and BP contributed to data acquisition, collection, and validation. SR contributed to health economic methodology and interpretation. LB, BFD, GR, GLC, PS, URP, CH, PK, REC, EJS, AMAl, HY, GSC, and DW contributed to clinical interpretation and critical manuscript revision. ASh supervised the study and provided critical intellectual input throughout manuscript development. All authors reviewed and approved the final manuscript and agreed to be accountable for the integrity of the work.

